# Comprehensive Evaluation of COVID-19 Patient Short- and Long-term Outcomes: Disparities in Healthcare Utilization and Post-Hospitalization Outcomes

**DOI:** 10.1101/2021.09.07.21263213

**Authors:** Stephen Salerno, Yuming Sun, Emily Morris, Xinwei He, Yajing Li, Ziyang Pan, Peisong Han, Jian Kang, Michael W. Sjoding, Yi Li

## Abstract

**Background:** Understanding risk factors for short- and long-term COVID-19 outcomes have implications for current guidelines and practice. We study whether early identified risk factors for COVID-19 persist one year later and through varying disease progression trajectories.

**Methods:** This was a retrospective study of 6,731 COVID-19 patients presenting to Michigan Medicine between March 10, 2020 and March 10, 2021. We describe disease progression trajectories from diagnosis to potential hospital admission, discharge, readmission, or death. Outcomes pertained to all patients: rate of medical encounters, hospitalization-free survival, and overall survival, and hospitalized patients: discharge versus in-hospital death and readmission. Risk factors included patient age, sex, race, body mass index, and 29 comorbidity conditions.

**Results:** Younger, non-Black patients utilized healthcare resources at higher rates, while older, male, and Black patients had higher rates of hospitalization and mortality. Diabetes with complications, coagulopathy, fluid and electrolyte disorders, and blood loss anemia were risk factors for these outcomes. Diabetes with complications, coagulopathy, fluid and electrolyte disorders, and blood loss were associated with lower discharge and higher inpatient mortality rates.

**Conclusions:** This study found differences in healthcare utilization and adverse COVID-19 outcomes, as well as differing risk factors for short- and long-term outcomes throughout disease progression. These findings may inform providers in emergency departments or critical care settings of treatment priorities, empower healthcare stakeholders with effective disease management strategies, and aid health policy makers in optimizing allocations of medical resources.

## Introduction

On March 10, 2020, the first confirmed cases of severe acute respiratory syndrome coronavirus 2 (SARS-CoV-2) were reported in the state of Michigan.[1] Since then, Southeast Michigan quickly evolved into an endemic center in the first wave of the pandemic, characterized by several densely populated urban areas, including Detroit.[2] Over the course of one year, the extent of the ensuing pandemic has changed drastically. More than 600,000 individuals in this country have died as a result of COVID-19,[3] with more than 21,000 from Michigan.[4, 5] The country was devastated with a healthcare crisis and economic wreckage,[6, 7] with medical resources depleted in endemic centers[8, 9] and roughly 20 million jobs lost nationwide.[10] More than 25% of American families currently experience financial hardship.[11, 12]

A growing body of literature has identified risk factors for COVID-19 outcomes, including older age,[13] male sex,[14] higher body mass index,[13] and comorbidity conditions such as cardiovascular disease,[15] diabetes,[16] chronic respiratory disease,[16, 17] hypertension,[18] and cancer.[19] Evidence of racial, ethnic,[20–23] and socioeconomic disparity[20, 22] in COVID-19 outcomes has further punctuated the clinical and broader societal impact of the pandemic.[24, 25] However, much of this early work that reported on the first wave of the pandemic was potentially limited by sample size or follow-up duration.[26–28] Further, most of this work relied on risk factors collected during the early phase of the pandemic.[29–31] As the effects of the pandemic have extended over the past year and into the foreseeable future, and the demographics of new cases are constantly changing,[32] it is pertinent to re-examine these risk factors for adverse outcomes in the context of new information.[33] Because of the limited scope of data, most previous work focused on one or two outcomes of COVID-19 patients,[13–16] lacking a comprehensive evaluation of disease progression. Indeed, owing to a longer follow-up available, especially among the earliest of cases in Southeast Michigan, it warranted studying outcomes that were less understood, such as healthcare utilization over time and competing hospitalization outcomes.

There is substantial interest to study COVID-19 outcomes in localized regions, such as Southeast Michigan, as it may lead to better allocation of healthcare resources and more effective preventive and disease management measures for those regions hit hardest by the pandemic.[20, 34, 35] One year after the first reported case in Michigan, this study sought to re-evaluate these proximate outcomes among COVID-19 patients in this early hotspot and examine the relationship between previously-identified risk factors and a variety of short- and long-term outcomes.

The University of Michigan (UM) Health System, referred to as Michigan Medicine hereafter, is one of the primary regional centers managing the care of COVID-19 patients [36, 37] and has created, maintained, and updated an electronic medical record (EMR) database for COVID-19 patients treated in its hospital system since the outbreak.[38] Access to this rich database enables us to conduct a comprehensive analysis of COVID-19 outcomes. In the following, we describe the disease progression trajectories of COVID-19 patients, from a positive test to potential hospital admissions, discharge, readmission post-discharge, or death. We then show modeling results for these outcomes on risk factors established in previous work.[13–19] We determine which of these risk factors persist as predictors for severe outcomes beyond the initial outbreak and throughout the past year as infections became more widespread. Our results have implications for current public health guidelines and policy, as COVID-19 has fundamentally changed the landscape of healthcare utilization.

## Materials and Methods

### Study Population

This was a retrospective study, approved by the UM Institutional Review Board, on multiple outcomes of COVID-19 patients and their associated risk factors. Included were COVID-19 patients who were treated at Michigan Medicine between March 10, 2020 and March 10, 2021. These patients were tested positive either at Michigan Medicine or elsewhere, with a positive test referring to a result of “detected,” “presumptive positive,” or “positive” obtained via reverse transcription polymerase chain reaction (PCR) tests on samples collected from sputum or from nasopharyngeal or oropharyngeal swabs (diagnosis code of U07.1 or U07.2). A small proportion of patients who transferred in and were tested elsewhere did not have dates or test results confirmed in their laboratory records, and, thus, had to be removed from our analysis. Our analyzable population was 6,731 patients with a positive COVID-19 diagnosis and available demographic and clinical data between March 10, 2020 and March 10, 2021 in the Michigan Medicine EMR. Their outcomes, demographic information, and clinical characteristics are described herein.

### Outcomes

Outcomes were categorized into three groups: those pertaining to all COVID-19 patients (1a-1c), those pertaining to hospitalized patients (2a-2c), and those for sensitivity analyses (3a-3b). They include:

- 1a. Rate of Medical Encounters: defined by each patient’s number of encounters with Michigan Medicine from COVID-19 diagnosis until the end of the study (March 10, 2021) divided by the number of days they were at risk.
- 1b. Hospitalization-Free Survival:[39, 40] measured as the time from COVID-19 diagnosis to first admission date or death, whichever came first, subject to censoring by the end of this study.
- 1c. Overall Survival: the time from diagnosis until death or administrative censoring at the end of the study.
- 2a-b. Discharge versus In-Hospital Death: the time from the admission date to (2a) discharge or (2b) in-hospital death, two competing short-term outcomes of hospitalization.
- 2c. Readmission: a binary variable indicating whether a patient was re-admitted at any point after discharge from their first hospitalization.
- 3a. Post-Admission Mortality: the time from patient’s first admission date post-diagnosis until death, possibly censored by the end of the study. This was to check the consistency of the results in 2a-b by incorporating any death, rather than only in-hospital death.
- 3b. COVID-Induced Hospitalizations/Readmissions: we restricted hospitalizations to COVID-specific admissions, and similarly, COVID-related readmissions following a COVID-19 hospitalization. This was to check the robustness of the results in 2a-c by only focusing on COVID-induced hospitalizations and readmissions.

### Demographic and Clinical Characteristics

We identified demographic data for all patients, including age, sex, race, and body mass index. Due to the homogeneity of the patient population at Michigan Medicine, patient’s self-identified race was classified into three groups: Black, non-Black, or Unknown. Similarly, due to few patients identifying as Hispanic or Latinx (< 5%), we excluded patient ethnicity as a potential covariate. Using available International Classification of Diseases, Tenth Revision (ICD-10) codes [41] (Supplement A, Table S1) from patient encounters, we constructed indicators for 29 of the 30 comorbidity conditions used in the Elixhauser comorbidity index,[42–44] the only exception being HIV/AIDS status due to its unavailability. Collectively, these comorbidity conditions were reported to be predictive of in-hospital and post-discharge mortality in numerous non-COVID [45–47] and COVID-19 settings.[15, 48–54]

### Statistical Analysis

We studied a natural disease progression by sequentially modeling the COVID-19 outcomes defined in above; see Figure 1 for a flow diagram depicting relationships between at-risk populations, patient outcomes, and corresponding models. First, to assess healthcare utilization among all COVID-positive patients, we modelled the associations of all potential risk factors with the rate of medical encounters in Poisson regression.[55] Patient who died before the end of the study were excluded from this model to mitigate survivorship bias. This was used as a proxy measure for healthcare utilization post-diagnosis, an outcome seldom investigated by the COVID-19 literature. We then studied the impact of risk factors on hospitalization-free survival and overall survival using Cox proportional hazards regression models.[56] Among hospitalized patients, we accounted for the competing relationship between two short-term outcomes of hospitalization, that is, discharge versus in hospital death, using the Fine-Gray subdistribution regression model;[57] see Supplement B for a more detailed description of the model. Finally, we modeled the odds of readmission among discharged patients using a logistic regression model.

**Figure 1:**
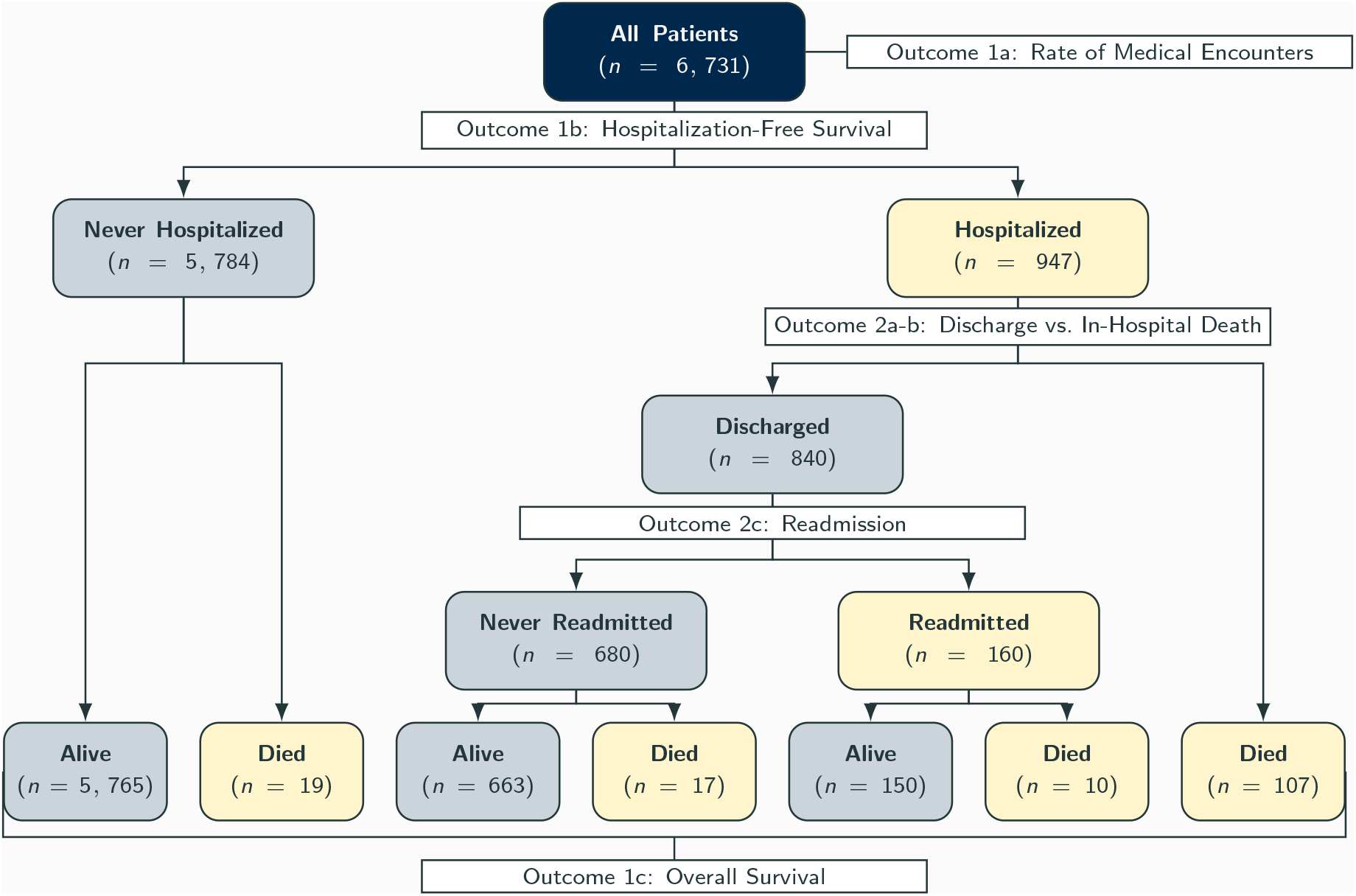
Diagram of disease progression for *N* = 6, 731 COVID-19 patients, and the relationship between the patient outcomes, patient sub-populations, and models in our analytic workflow.

## Results

### Patient Outcomes and Characteristics

Among 6,731 analyzable patients with a positive COVID-19 diagnosis, 947 (14%) were hospitalized at or after diagnosis and 153 died (2%) during the follow-up period. Among those hospitalized, 840 (89%) were discharged and 160 (17%) were subsequently readmitted. We observed 134 deaths among hospitalized patients: 107 (81%) in-hospital deaths, i.e., deaths before discharge from their first (index) hospitalization, 17 (12%) after their index discharge, and 10 (7%) after readmission; see Figure 1.

On average, these 6,731 COVID-19 patients were 44 years old and majority female (56%), with an over-representation of Black patients (15%) as compared to the general population surrounding Michigan Medicine. There was a high proportion of patients with cardiac arrhythmias (27%), hypertension (32% uncomplicated, 9% complicated), chronic pulmonary disease (26%), obesity (28%), and fluid and electrolyte disorders (20%; Table 1). Of note, a total of 539 (8%) patients did not disclose their race; Table S2 (Supplement A) shows they were, in general, much younger and healthier than those who identified their race.

**Table 1:** Descriptive characteristics for COVID-19 patients treated at Michigan Medicine between March 10, 2020 and March 10, 2021. Summary statistics are for the overall population, those admitted for an inpatient stay during the follow-up period, and those readmitted post-discharge.

| Characteristic | All Patients<br>N = 6,731<sup>a</sup> | Hospitalized Patients<br>N = 947<sup>a</sup> | Readmitted Patients<br>N = 160<sup>a</sup> |
| --- | --- | --- | --- |
| Age (years) | 44 (24, 60) | 60 (46, 71) | 61 (40, 73) |
| Sex |  |  |  |
| Female | 3,706 (56%) | 430 (45%) | 59 (37%) |
| Male | 2,952 (44%) | 517 (55%) | 101 (63%) |
| Race |  |  |  |
| White | 4,560 (68%) | 518 (55%) | 99 (62%) |
| Black | 1,025 (15%) | 305 (32%) | 49 (31%) |
| Asian | 252 (3.7%) | 29 (3.1%) | 5 (3.1%) |
| Multicultural/Other | 315 (4.7%) | 43 (4.5%) | 6 (3.8%) |
| Unknown | 579 (8.6%) | 52 (5.5%) | 1 (0.6%) |
| Ethnicity |  |  |  |
| Non-Hispanic or Latino | 5,741 (85%) | 867 (92%) | 156 (98%) |
| Hispanic or Latino | 267 (4.0%) | 25 (2.6%) | 3 (1.9%) |
| Unknown | 579 (8.6%) | 52 (5.5%) | 1 (0.6%) |
| Body Mass Index (kg/m2) | 30 (28, 30) | 30 (26, 36) | 27 (24, 34) |
| Congestive Heart Failure | 582 (8.6%) | 235 (25%) | 54 (34%) |
| Cardiac Arrhythmias | 1,797 (27%) | 587 (62%) | 133 (83%) |
| Valvular Disease | 391 (5.8%) | 111 (12%) | 34 (21%) |
| Pulmonary Circulation Disorders | 399 (5.9%) | 187 (20%) | 48 (30%) |
| Peripheral Vascular Disorders | 543 (8.1%) | 193 (20%) | 61 (38%) |
| Hypertension, Uncomplicated | 2,139 (32%) | 636 (67%) | 123 (77%) |
| Hypertension, Complicated | 627 (9.3%) | 336 (35%) | 74 (46%) |
| Paralysis | 138 (2.1%) | 61 (6.4%) | 23 (14%) |
| Other Neurological Disorders | 616 (9.2%) | 246 (26%) | 66 (41%) |
| Chronic Pulmonary Disease | 1,730 (26%) | 370 (39%) | 72 (45%) |
| Diabetes, Uncomplicated | 1,049 (16%) | 383 (40%) | 71 (44%) |
| Diabetes, Complicated | 743 (11%) | 330 (35%) | 64 (40%) |
| Hypothyroidism | 715 (11%) | 151 (16%) | 34 (21%) |
| Renal Failure | 744 (11%) | 334 (35%) | 72 (45%) |
| Liver Disease | 710 (11%) | 186 (20%) | 52 (32%) |
| Lymphoma | 119 (1.8%) | 32 (3.4%) | 5 (3.1%) |
| Metastatic Cancer | 595 (8.8%) | 139 (15%) | 48 (30%) |
| Solid Tumor without Metastasis | 674 (10%) | 161 (17%) | 47 (29%) |
| Rheumatoid Arthritis/Collagen Vascular Diseases | 536 (8.0%) | 98 (10%) | 27 (17%) |
| Coagulopathy | 662 (9.8%) | 298 (31%) | 80 (50%) |
| Obesity | 1,880 (28%) | 443 (47%) | 71 (44%) |
| Weight Loss | 632 (9.4%) | 232 (24%) | 74 (46%) |
| Fluid and Electrolyte Disorders | 1,349 (20%) | 632 (67%) | 137 (86%) |
| Blood Loss Anemia | 276 (4.1%) | 121 (13%) | 45 (28%) |
| Deficiency Anemia | 660 (9.8%) | 183 (19%) | 55 (34%) |
| Alcohol Abuse | 285 (4.2%) | 55 (5.8%) | 14 (8.8%) |
| Drug Abuse | 336 (5.0%) | 101 (11%) | 34 (21%) |
| Psychoses | 201 (3.0%) | 84 (8.9%) | 23 (14%) |
| Depression | 1,835 (27%) | 369 (39%) | 99 (62%) |

<sup>a</sup>Median (IQR); n (%)

Restricting to the 947 hospitalized patients, we observed a much 192 higher average age of 60 years old and a higher proportion of male (55%) and Black patients (32%), as well as a consistently higher comorbidity burden. Further restricting to the 160 patients with multiple admissions, this trend persisted, with an average age of 61 years old, 83% of patients diagnosed with cardiac arrhythmias, 77% with hypertension, 86% with fluid and electrolyte disorders, and 62% with depressive symptoms.

### Risk Factors for Outcomes among All COVID-19 Patients

In examining medical utilization among all COVID-19 patients, we found that younger (per 10 years; Incidence Rate Ratio (IRR): 0.97; 95% Confidence Interval (CI): 0.97-0.98) and non-Black (1.02; 1.00-1.03) patients had higher hospital utilization rates, and that 22 of 29 comorbidity conditions were associated with significantly higher hospital utilization rates (Table 2). Cardiac arrythmias (1.38; 1.36-1.39), fluid and electrolyte disorders (1.50; 1.49-1.52), and blood loss anemia (1.37; 1.35-1.39) were among the most significant comorbidity conditions for increased medical utilization.

**Table 2:** Adjusted associations of patient demographic and clinical risk factors with COVID-207 19 outcomes among all COVID-19 patients, based on (a) Poisson regression for the number of encounters each patient had with Michigan Medicine; (b) Cox proportional hazards regression for hospitalization-free survival; (c) Cox proportional hazards regression for overall survival.

| Characteristic | Number of Encounters<br>IRR (95% CI)<sup>a</sup> | Overall-Mortality<br>HR (95% CI)<sup>a</sup> |
| --- | --- | --- |
| Age (per 10 years) | **0.97 (0.97, 0.98)<sup>b</sup>** | **1.69 (1.47, 1.93)** |
| Sex |  |  |
| Female | — | — |
| Male | 1.00 (0.99, 1.01) | 1.34 (0.93, 1.92) |
| Race |  |  |
| Black | — | — |
| Non-Black | **1.02 (1.00, 1.03)** | **0.66 (0.45, 0.98)** |
| Unknown | **0.73 (0.71, 0.76)** | **2.57 (1.44, 4.59)** |
| Body Mass Index (kg/m<sup>2</sup>) | **0.99 (0.99, 0.99)** | 0.98 (0.96, 1.01) |
| Congestive Heart Failure | **1.02 (1.00, 1.03)** | 1.13 (0.75, 1.69) |
| Cardiac Arrhythmias | **1.38 (1.36, 1.39)** | **1.61 (1.04, 2.50)** |
| Valvular Disease | **0.93 (0.92, 0.95)** | 0.91 (0.58, 1.43) |
| Pulmonary Circulation Disorders | **1.21 (1.20, 1.23)** | 1.09 (0.73, 1.64) |
| Peripheral Vascular Disorders | **1.10 (1.08, 1.11)** | 0.80 (0.52, 1.21) |
| Hypertension, Uncomplicated | **1.21 (1.19, 1.22)** | 0.72 (0.45, 1.16) |
| Hypertension, Complicated | **1.06 (1.04, 1.08)** | 1.51 (0.95, 2.41) |
| Paralysis | **1.45 (1.42, 1.48)** | 1.37 (0.71, 2.68) |
| Other Neurological Disorders | **1.05 (1.04, 1.07)** | **1.49 (1.03, 2.14)** |
| Chronic Pulmonary Disease | **1.05 (1.04, 1.06)** | **1.70 (1.19, 2.44)** |
| Diabetes, Uncomplicated | 0.99 (0.97, 1.00) | 0.88 (0.54, 1.44) |
| Diabetes, Complicated | **1.14 (1.12, 1.16)** | **2.05 (1.24, 3.39)** |
| Hypothyroidism | **1.12 (1.11, 1.14)** | 0.65 (0.40, 1.05) |
| Renal Failure | **0.97 (0.95, 0.98)** | 1.04 (0.68, 1.57) |
| Liver Disease | **1.08 (1.07, 1.10)** | 0.93 (0.62, 1.40) |
| Lymphoma | **1.19 (1.16, 1.22)** | 1.39 (0.68, 2.86) |
| Metastatic Cancer | **1.22 (1.20, 1.24)** | 1.02 (0.60, 1.73) |
| Solid Tumor without Metastasis | **1.14 (1.13, 1.16)** | 0.88 (0.54, 1.43) |
| Rheumatoid Arthritis/Collagen Vascular Diseases | **1.21 (1.19, 1.22)** | 0.91 (0.53, 1.55) |
| Coagulopathy | **1.26 (1.24, 1.28)** | **2.02 (1.39, 2.92)** |
| Obesity | **1.34 (1.32, 1.36)** | 1.28 (0.85, 1.95) |
| Weight Loss | **1.26 (1.24, 1.27)** | 0.85 (0.57, 1.27) |
| Fluid and Electrolyte Disorders | **1.50 (1.49, 1.52)** | **5.50 (3.27, 9.23)** |
| Blood Loss Anemia | **1.37 (1.35, 1.39)** | **2.85 (1.84, 4.40)** |
| Deficiency Anemia | **1.06 (1.05, 1.08)** | **0.44 (0.28, 0.69)** |
| Alcohol Abuse | **0.94 (0.92, 0.96)** | 0.71 (0.37, 1.37) |
| Drug Abuse | **0.97 (0.95, 0.98)** | 0.95 (0.52, 1.74) |
| Psychoses | 0.99 (0.97, 1.01) | 1.06 (0.63, 1.80) |
| Depression | **1.33 (1.32, 1.35)** | **0.60 (0.41, 0.89)** |

<sup>a</sup>IRR = Incidence Rate Ratio, CI = Confidence Interval, HR = Hazard Ratio
<sup>b</sup>Bold values indicate statistical significance

We further identified several risk factors that persisted as significant predictors of acute outcomes, namely hospitalization-free survival and overall survival. Older (per 10 years; Hazard Ratio (HR): 1.23; 95% CI: 1.18-1.28), male (1.34; 1.16-1.54), and Black patients (1.89; 95% CI: 1.61-2.17) had higher hospitalization hazards. Additionally, ten comorbidity conditions were associated with higher hazards of hospitalization: cardiac arrhythmias (1.77; 1.51-2.07), pulmonary circulation disorders (1.27; 1.05-1.53), hypertension with (1.21; 1.01-1.45) and without complications (1.49; 1.22-1.83), other neurological disorders (1.35; 1.14-1.60), diabetes with complications (1.28; 1.02-1.60), coagulopathy (1.87; 1.60-2.20), weight loss (1.22; 1.03-1.44), fluid and electrolyte disorders (4.48; 3.78-5.30), and blood loss anemia (1.32; 1.06-1.64), while five were associated with lower hospitalizations hazards: valvular disease (0.71; 0.57-0.89), liver disease (0.79; 0.67-0.95), rheumatoid arthritis/collagen vascular diseases (0.80; 0.64-0.99), deficiency anemia (0.70; 0.58-0.84), and alcohol abuse (0.62; 0.46-0.83; see Supplement C, Table S3). Considering overall survival, older (per 10 years; HR: 1.69; 95% CI: 1.47-1.93) and Black patients (1.52; 1.02-2.22), as well as those with cardiac arrythmias (1.61; 1.04-2.50), chronic pulmonary disease (1.70; 1.19-2.44), diabetes with complications (2.05; 1.24-3.39), coagulopathy (2.02; 1.39-2.92), fluid and electrolyte disorders (5.50; 3.27-9.23), and blood loss anemia (2.85; 1.84-4.40), had significantly higher mortality (Table 2).

### Risk Factors for Post-Hospitalization Outcomes

Among patients hospitalized after COVID-19 diagnosis, we modeled in-hospital death versus discharge, two competing hospitalization outcomes, using the Fine-Gray subdistribution hazards model (Table 3). Older age (per 10 years) was associated with a 9% lower discharge rate (Subdistribution Hazard Ratio (SHR): 0.91; 95% CI: 0.86-0.95) and a 50% higher in-hospital mortality rate (SHR: 1.50; 95% CI: 1.27-1.77) post-diagnosis. Diabetes with complications (Discharge: 0.71; 0.57-0.88; Mortality: 1.82; 1.00-3.33), coagulopathy (Discharge: 0.71; 0.61-0.84; Mortality: 1.72; 1.15-2.56), fluid and electrolyte disorders (Discharge: 0.70; 0.60-0.82; Mortality: 2.57; 1.30-5.07), and blood loss anemia (Discharge: 0.67; 0.53-0.85; Mortality: 2.68; 1.66-4.32) were associated with lower discharge and higher inpatient mortality rates. Additionally, male sex (0.82; 0.71-0.95), cardiac arrhythmias (0.85; 0.73-0.99), and other neurological disorders (0.73; 0.61-0.87) were associated with lower discharge rates, while chronic pulmonary disease (1.69; 1.12-2.55) was associated with higher inpatient mortality. In contrast, solid tumor cancers (1.27; 1.01-1.59), rheumatoid arthritis/collagen vascular diseases (1.34; 1.07-1.68), and drug abuse (1.31; 1.03-1.67) were associated with higher discharge rates, while weight loss (0.53; 0.31-0.90), deficiency anemia (0.44; 0.24-0.82), and depression (0.57; 0.34-0.97) were associated with lower inpatient mortality rates (Table 3). In a sensitivity analysis, we restricted our competing risks analysis to patients hospitalized directly due to COVID-19 and found similar patterns of associations (Supplement D, Table S5).

**Table 3:** Adjusted associations of patient demographic and clinical risk factors with COVID-19 outcomes among hospitalized COVID-19 patients, based on Fine-Gray subdistribution hazards regression for (a) discharge post-admission and (b) in-hospital death post-admission and logistic regression for (c) whether the patient was readmitted at any point post-index discharge.

| Characteristic | Discharge<br>SHR (95% CI)<sup>a</sup> | In-Hospital Death<br>SHR (95% CI)<sup>a</sup> |
| --- | --- | --- |
| Age (per 10 years) | **0.91 (0.86, 0.95)<sup>b</sup>** | **1.50 (1.27, 1.77)** |
| Sex |  |  |
| Female | — | — |
| Male | **0.82 (0.71, 0.95)** | 1.38 (0.85, 2.23) |
| Race |  |  |
| Black | — | — |
| Non-Black | 1.12 (0.96, 1.30) | 0.69 (0.43, 1.12) |
| Unknown | **0.47 (0.31, 0.69)** | **2.42 (1.22, 4.77)** |
| Body Mass Index (kg/m<sup>2</sup>) | 1.00 (0.99, 1.01) | 0.97 (0.94, 1.01) |
| Congestive Heart Failure | 0.88 (0.72, 1.09) | 1.31 (0.80, 2.16) |
| Cardiac Arrhythmias | **0.85 (0.73, 0.99)** | 1.42 (0.83, 2.44) |
| Valvular Disease | 1.17 (0.91, 1.51) | 0.99 (0.55, 1.79) |
| Pulmonary Circulation Disorders | 0.84 (0.70, 1.01) | 1.02 (0.64, 1.62) |
| Peripheral Vascular Disorders | 1.20 (0.98, 1.46) | 0.75 (0.44, 1.26) |
| Hypertension, Uncomplicated | 1.01 (0.85, 1.19) | 0.72 (0.43, 1.20) |
| Hypertension, Complicated | 1.18 (0.96, 1.46) | 0.86 (0.48, 1.52) |
| Paralysis | 1.12 (0.86, 1.45) | 0.79 (0.30, 2.08) |
| Other Neurological Disorders | **0.73 (0.61, 0.87)** | 1.44 (0.94, 2.19) |
| Chronic Pulmonary Disease | 0.87 (0.75, 1.01) | **1.70 (1.13, 2.55)** |
| Diabetes, Uncomplicated | 1.09 (0.89, 1.33) | 0.91 (0.51, 1.62) |
| Diabetes, Complicated | **0.71 (0.57, 0.88)** | **1.82 (1.00, 3.33)** |
| Hypothyroidism | 1.05 (0.86, 1.28) | 0.65 (0.32, 1.30) |
| Renal Failure | 0.89 (0.73, 1.08) | 1.12 (0.68, 1.86) |
| Liver Disease | 0.98 (0.81, 1.18) | 1.21 (0.75, 1.96) |
| Lymphoma | 0.69 (0.43, 1.11) | 1.63 (0.66, 4.03) |
| Metastatic Cancer | 1.18 (0.91, 1.53) | 0.99 (0.49, 1.97) |
| Solid Tumor without Metastasis | **1.27 (1.01, 1.59)** | 0.66 (0.34, 1.26) |
| Rheumatoid Arthritis/Collagen Vascular Diseases | **1.34 (1.07, 1.68)** | 0.80 (0.36, 1.77) |
| Coagulopathy | **0.71 (0.61, 0.84)** | **1.61 (1.07, 2.42)** |
| Obesity | 0.99 (0.83, 1.17) | 1.53 (0.94, 2.49) |
| Weight Loss | 0.90 (0.77, 1.05) | **0.53 (0.31, 0.90)** |
| Fluid and Electrolyte Disorders | **0.70 (0.60, 0.82)** | **2.57 (1.30, 5.07)** |
| Blood Loss Anemia | **0.67 (0.53, 0.85)** | **2.68 (1.66, 4.32)** |
| Deficiency Anemia | 1.09 (0.90, 1.33) | **0.44 (0.24, 0.82)** |
| Alcohol Abuse | 1.18 (0.86, 1.61) | 0.61 (0.27, 1.40) |
| Drug Abuse | **1.31 (1.03, 1.67)** | 0.81 (0.34, 1.92) |
| Psychoses | 0.94 (0.71, 1.23) | 1.12 (0.58, 2.15) |
| Depression | 1.06 (0.92, 1.23) | **0.57 (0.34, 0.97)** |

<sup>a</sup>SHR = Subdistribution Hazard Ratio, CI = Confidence Interval
<sup>b</sup>Bold values indicate statistical significance

In a second sensitivity analysis, we considered risk factors leading to post-admission mortality among hospitalized patients. Post-admission survival was defined as the time lag between the patient’s first admission date post-diagnosis and death, subject to administrative censoring on March 10, 2021. Figure 2 plots Kaplan-Meier curves for post-admission survival, stratified by age quartiles, chronic pulmonary disease, coagulopathy, fluid and electrolyte disorders, blood loss anemia, and depression, which were significantly associated with post-admission mortality using univariate log-rank tests.

**Figure 2:**
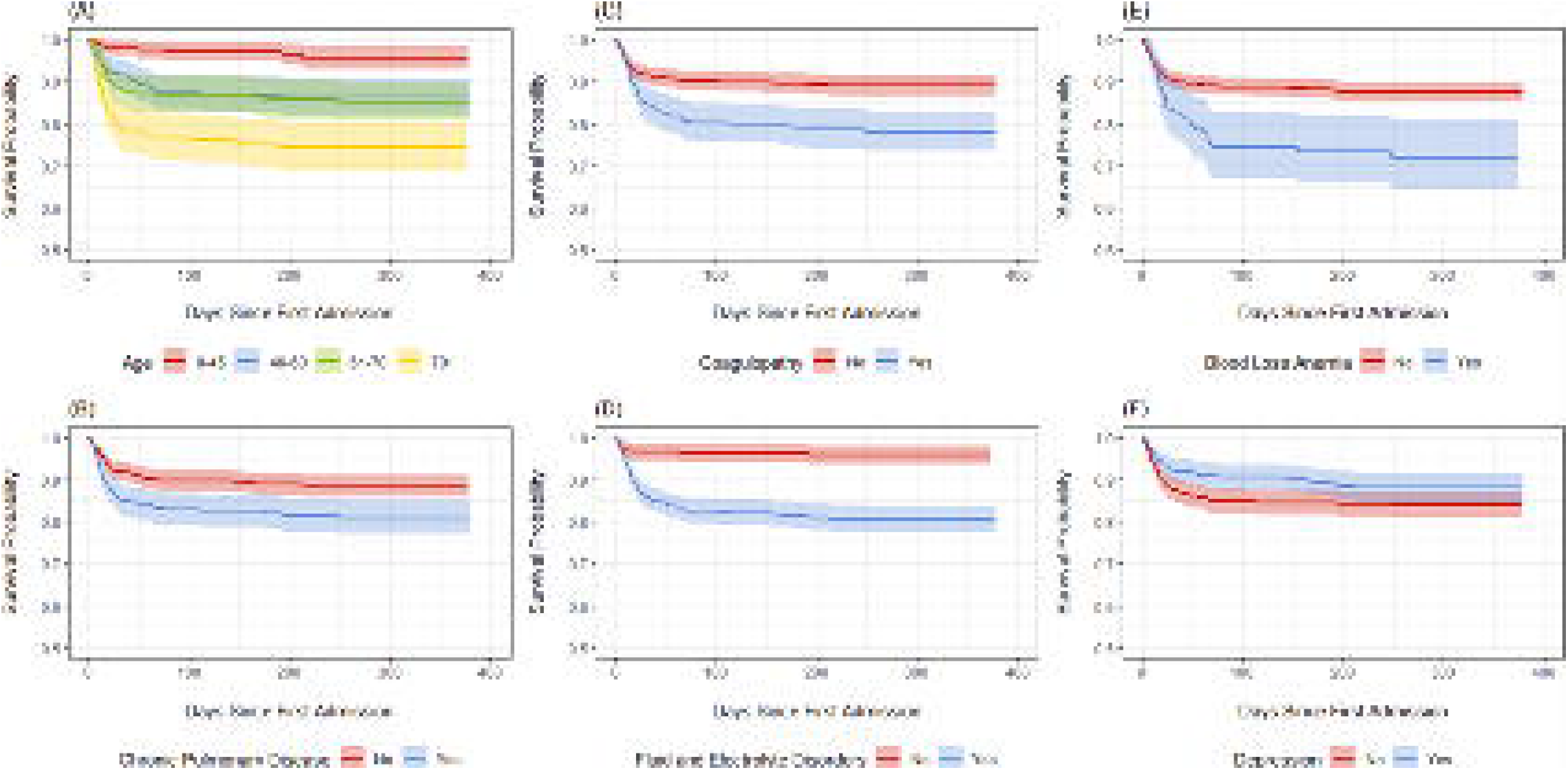
Kaplan-Meier curves for post-admission mortality among hospitalized COVID-19 patients, stratified by (A) age quartiles, (B) chronic pulmonary disease, (C) coagulopathy, (D) fluid and electrolyte disorders, (E) blood loss anemia, and (F) depression.

We then used a Cox regression model to detect risk factors associated with post-admission mortality among hospitalized patients, adjusting for all other factors. Differing from our competing risks analysis, this model defines the event of interest to be death at any point following an inpatient stay, including among patients who died post-discharge. We found that older age (per 10 years) was associated with 45% higher post-admission mortality hazard (HR: 1.45; 95% CI: 1.25-1.68) and that chronic pulmonary disease (1.63; 1.12-2.37), coagulopathy (1.54; 1.05-2.26), fluid and electrolyte disorders (2.90; 1.57-3.69), and blood loss anemia (2.33; 1.47-3.69) were significantly associated with higher mortality rates, while hypertension without complications (0.61; 0.38-1.00) and depression (0.65; 0.42-1.00) were associated with lower mortality rates (Table 4).

**Table 4:** Adjusted associations of patient demographic and clinical risk factors with COVID-19 outcomes among hospitalized COVID-19 patients, based on a Cox proportional hazards regression for time to any death post-admission.

| Characteristic | Post-Admission Mortality<br>HR (95% CI)<sup>1</sup> |
| --- | --- |
| Age (per 10 years) | **1.45 (1.25, 1.68)** |
| Sex |  |
| Female | — |
| Male | 1.28 (0.86, 1.90) |
| Race |  |
| Black | — |
| Non-Black | 0.90 (0.59, 1.38) |
| Unknown | **3.04 (1.67, 5.56)** |
| Body Mass Index (kg/m<sup>2</sup>) | 0.98 (0.95, 1.01) |
| Congestive Heart Failure | 1.21 (0.77, 1.89) |
| Cardiac Arrhythmias | 1.51 (0.93, 2.45) |
| Valvular Disease | 0.91 (0.55, 1.51) |
| Pulmonary Circulation Disorders | 1.04 (0.68, 1.58) |
| Peripheral Vascular Disorders | 0.95 (0.60, 1.50) |
| Hypertension, Uncomplicated | **0.61 (0.38, 1.00)** |
| Hypertension, Complicated | 1.37 (0.81, 2.32) |
| Paralysis | 0.93 (0.44, 1.94) |
| Other Neurological Disorders | 1.29 (0.87, 1.90) |
| Chronic Pulmonary Disease | **1.63 (1.12, 2.37)** |
| Diabetes, Uncomplicated | 0.94 (0.56, 1.57) |
| Diabetes, Complicated | 1.60 (0.96, 2.67) |
| Hypothyroidism | 0.76 (0.45, 1.28) |
| Renal Failure | 0.95 (0.60, 1.50) |
| Liver Disease | 0.93 (0.59, 1.47) |
| Lymphoma | 1.42 (0.59, 3.42) |
| Metastatic Cancer | 1.30 (0.70, 2.42) |
| Solid Tumor without Metastasis | 0.76 (0.42, 1.37) |
| Rheumatoid Arthritis/Collagen Vascular Diseases | 0.56 (0.26, 1.18) |
| Coagulopathy | **1.54 (1.05, 2.26)** |
| Obesity | 1.33 (0.85, 2.09) |
| Weight Loss | 0.86 (0.56, 1.32) |
| Fluid and Electrolyte Disorders | **2.90 (1.57, 5.34)** |
| Blood Loss Anemia | **2.33 (1.47, 3.69)** |
| Deficiency Anemia | **0.56 (0.34, 0.92)** |
| Alcohol Abuse | 0.73 (0.33, 1.63) |
| Drug Abuse | 0.62 (0.29, 1.34) |
| Psychoses | 1.16 (0.65, 2.09) |
| Depression | **0.65 (0.42, 1.00)** |

<sup>1</sup>HR = Subdistribution Hazard Ratio, CI = Confidence Interval
<sup>2</sup>Bold values indicate statistical significance

Studying the risk factors for readmission post-discharge, we found 256 that cardiac arrhythmias (Odds Ratio (OR): 1.85; 95% CI: 1.12-3.13), peripheral vascular disorders (1.66; 1.01-2.71), weight loss (1.69; 1.10-2.59), fluid and electrolyte disorders (1.84; 1.08, 3.22), blood loss anemia (1.81; 1.06-3.08), and depression (2.20; 1.42-3.42) were significantly associated with higher odds of readmission, while older age (per ten years) was associated with a 16% lower odds of readmission (0.84; 0.74-0.95) and chronic pulmonary disease had lower odds of readmission (0.63; 0.40-0.98; Supplement C, Table S4). These results were qualitatively consistent with a sensitivity analysis which restricted index hospitalizations and subsequent readmissions to instances where COVID-19 was the reason for admission (Supplement D, Table S5).

## Discussion

Leveraging EMR data from a regional medical center managing COVID-19, this study enriches the literature with a large COVID-19 positive patient cohort and a longer follow-up period to observe evolving patient outcomes. Differing outcomes associated with demographic characteristics such as age, sex, and race, which have become particularly salient,[20, 22, 37, 58-60] were corroborated in this study. Notably, after adjusting for all other risk factors, Black patients had lower healthcare utilization rates, but higher hospitalization and mortality rates. Previous work has similarly noted these differences in adverse COVID-19 outcomes between Black and non-Black patients.[61-65] Higher comorbidity burden was also shown to be associated with these outcomes, particularly among patients with chronic pulmonary disease, diabetes, and cardiac complications.[15, 48–51]

With regards to less-studied post-hospitalization outcomes, fluid and electrolyte disorders and blood loss anemia were associated with higher readmission and higher post-hospitalization mortality rates. While mechanisms of these associations are unknown, one potential explanation is syndrome of inappropriate antidiuretic hormone (SIADH) secretion. Hyponatremia and SIADH have a known association with severe COVID-19 infection, as reported previously.[66–68] However, patients in our study population who were broadly indicated for fluid and electrolyte disorders have imbalances that span the range of sodium and potassium, acidosis, alkalosis, and volume depletion which coincide with each other and worsened COVID-19 outcomes.[69-71] Additionally, blood loss anemia may be aggravated by repeated blood tests, particularly, in those who developed pneumonia and acute respiratory distress syndrome (ARDS), or due to gastrointestinal bleeding.[72-80] Lastly, depression was found to be associated with higher odds of readmission, but lower mortality rates. It is being shown that patients who experienced severe COVID-19 and had lasting long-term morbidity were more likely to develop depression or other psychiatric disorder,[81] and thus might be readmitted due to the residual effects of COVID-19.[82]

Patient age and pre-existing chronic pulmonary disease were associated with lower odds of readmission, but higher mortality rates, suggesting a greater risk of in-hospital mortality for these severe risk factors.[53, 83-86] Cardiac arrhythmias, peripheral vascular disorders, and coagulopathy were also associated with higher readmissions and post-hospitalization mortality. Cardiac complications among severe COVID-19 cases have been shown to include myocardial injury, heart failure, and sudden cardiac arrest.[87–90] Such involvement poses potential implications for monitoring patient prognosis during an inpatient stay, as well as long-term surveillance in recovered cases. Concerning risk factors associated with the two competing shorter-term hospitalization outcomes, discharge and in-hospital death, we identified that older age, diabetes with complications, coagulopathy, fluid and electrolyte disorders, and blood loss anemia were associated with lower rates of discharge and higher rates of in-hospital death. The findings corroborate previous results for in-hospital mortality, which did not take into account patient discharge as a competing event.[91–96] To our knowledge, we are among the first few to consider discharge versus in-hospital mortality among COVID-19 patients under a competing risk regression framework.[57] Ignoring their competing relationship can lead to biased estimates of effects of risk factors for either event.[97] A recent study[98] investigated COVID-19 hospitalization outcomes in a similar manner, reporting a subdistribution hazard ratio for patient age, adjusting for sex. This result, in a smaller, localized international cohort, is consistent with our findings on age.

Lastly, we failed to detect significant associations between risk factors such as obesity and renal failure, which have been studied extensively in the COVID-19 literature. While obesity, being pro-inflammatory, is a well-established risk factor for worsened outcomes in COVID-19 patients,[99-102] we failed to detect a statistically significant obesity effect in this subset of patients. Similarly, renal involvement with COVID-19 is well-studied, though multifactorial in nature.[103-106] We believe this possibly due to (1) potential collinearity with more down-stream risk factors included in our models and (2) a lack of statistical power to detect these effects, as the directions of several effects were consistent with previously established associations with obesity and renal failure, though not statistically significant. As a sensitivity check, we fit univariate models for each of the comorbidity conditions, adjusted for patient demographics (age, sex, and race). These results show significant associations, marginally, for these risk factors (see Supplement E).

### Limitations

First, as a small number of patients who transferred in from other institutions did not have medical history data, we had to remove them from analysis, though their impacts on our results were limited. Second, as this study was based exclusively on patients of Michigan Medicine, there may be biases in the patient mix, affecting the generalizability to more diverse populations or other geographic areas. On the other hand, these patients did offer an opportunity to study COVID-19 outcomes in a local region that had been severely impacted by the pandemic. Third, this was a retrospective study of an existing EMR database. As such, we are limited in our ability to draw causal interpretations from these results. In addition, due to the nature of EMR data, there is always the possibility for misclassification bias and/or inaccurate data entry. Lastly, future work, with longer follow-up, should focus on the residual impact of COVID-19 among recovered patients to elucidate the effect of lasting symptoms and acquired comorbidity burden on long-term quality of life and mortality.

## Conclusions

Several lessons can be learned with our analysis. First, there exist socio-demographic inequalities in healthcare access and COVID-19 outcomes among COVID-19 patients; after adjusting for all comorbid and other demographic conditions, being Black was associated higher hospitalization and mortality rates. Second, chronic pulmonary disease, diabetes, and cardiac complications were among the most significant risk factors for these outcomes, suggesting more targeted screening among at-risk patients. Third, among hospitalized COVID-19 patients, poorer outcomes may be disproportionately associated with additional risk factors, namely cardiac complications such as arrhythmias and coagulopathy, as well as other comorbid conditions such as peripheral vascular disorders, fluid and electrolyte disorders, blood loss anemia, and abnormal weight loss. Therefore, increased testing and vaccination efforts in these particularly susceptible populations are necessary to prevent residual disease burden. Lastly, these analysis results may inform providers in emergency departments or critical care settings of treatment priorities, empower healthcare stakeholders with effective disease management strategies, and aid health policy makers in optimizing allocations of medical resources.

## Data Availability

Data cannot be shared publicly due to patient confidentiality. The data underlying the results presented in the study are available from University of Michigan Data Office for Clinical & Translational Research for researchers who meet the criteria for access to confidential data.

## Acknowledgements

We thank Dr. Brahmajee Nallamothu for leading the development and curation of DataDirect, a newly launched, GPU-based analytics platform through the Michigan Medicine Precision Health Initiative. We are also grateful to Anisa Driscoll and Kate Smith for their continued analytical support with respect to database management, data processing, and use of the DataDirect platform.

## Supporting Information Captions

A. Comorbidity Conditions and Additional Comparisons Between Race
B. Fine-Gray Model for Competing Risks
C. Sensitivity Analysis

## Notes

### Competing Interest Statement

The authors have declared no competing interest.

### Funding Statement

YL is supported by the University of Michigan Precision Health Initiative (https://precisionhealth.umich.edu/) and National Institutes of Health - National Cancer Institute (https://www.cancer.gov/) grant R01CA249096. The funders had no role in study design, data collection and analysis, decision to publish, or preparation of the manuscript.

### Author Declarations

Study protocols were reviewed and approved by the University of Michigan Medical School Institutional Review Board (IRB ID HUM00192931).

