## Supplement for "Comprehensive Evaluation of COVID-19 Patient Short- and Long-term Outcomes: Disparities in Healthcare Utilization and Post-Hospitalization Outcomes"

Supplementary Materials

The following document provides supporting information for the manuscript: *Comprehensive Evaluation of COVID-19 Patient Short- and Long-term Outcomes: Disparities in Healthcare Utilization and Post-Hospitalization Outcomes*.

### B. Additional Information on Covariate Data

#### A.1. Definition of Comorbidity Conditions

Table S1 lists the 29 comorbidity conditions considered as risk factors in this analysis and the corresponding ICD-10 codes used to define them. Each comorbidity was coded as a binary indicator, flagging whether a patient carried any ICD-10 code associated with the condition between March 10, 2020 - March 10, 2021.

Table S1: Elixhauser comorbidity conditions and associated ICD-10 codes

| **Comorbidity Condition** | **ICD-10 Codes** |
| --- | --- |
| Congestive Heart Failure | I09.9, I11.0, I13.0, I13.2, I25.5, I42.0, I42.5-I42.9, I43.x, I50.x, P29.0 |
| Cardiac Arrhythmias | I44.1-I44.3, I45.6, I45.9, I47.x-I49.x, R00.0, R00.1, R00.8, T82.1, Z45.0, Z95.0 |
| Valvular Disease | A52.0, I05.x-I08.x, I09.1, I09.8, I34.x-I39.x, Q23.0-Q23.3, Z95.2- Z95.4 |
| Pulmonary Circulation Disorders | I26.x, I27.x, I28.0, I28.8, I28.9 |
| Peripheral Vascular Disorders | I70.x, I71.x, I73.1, I73.8, I73.9, I77.1, I79.0, I79.2, K55.1, K55.8, K55.9, Z95.8, Z95.9 |
| Hypertension, Uncomplicated | I10.x |
| Hypertension, Complicated | I11.x-I13.x, I15.x |
| Paralysis | G04.1, G11.4, G80.1, G80.2, G81.x, G82.x, G83.0, G83.4, G83.9 |
| Other Neurological Disorders | G10.x-G13.x, G20.x-G22.x, G25.4, G25.5, G31.2, G31.8, G31.9, G32.x, G35.x-G37.x, G40.x, G41.x, G93.1, G93.4, R47.0, R56.x |
| Chronic Pulmonary Disease | I27.8, I27.9, J40.x-J47.x, J60.x-J67.x, J68.4, J70.1, J70.3 |
| Diabetes, Uncomplicated | E10.0, E10.1, E10.9, E11.0, E11.1, E11.9, E12.0, E12.1, E12.9, E13.0, E13.1, E13.9, E14.0, E14.1, E14.9 |
| Diabetes, Complicated | E10.2-E10.8, E11.2-E11.8, E12.2-E12.8, E13.2-E13.8, E14.2-E14.8 |
| Hypothyroidism | E00.x-E03.x, E89.0 |
| Renal Failure | I12.0, I13.1, N18.x, N19.x, N25.0, Z49.0-Z49.2, Z94.0, Z99.2 |
| Liver Disease | B18.x, I85.x, I86.4, I98.2, K70.x, K71.1, K71.3-K71.5, K71.7, K72.x-K74.x, K76.0, K76.2-K76.9, Z94.4 |
| Lymphoma | C81.x-C85.x, C88.x, C96.x, C90.0, C90.2 |
| Metastatic Cancer | C77.x-C80.x |
| Solid Tumor, Without Metastasis | C00.x-C26.x, C30.x-C34.x, C37.x-C41.x, C43.x, C45.x-C58.x, C60.x-C76.x, C97.x |
| RA/Collagen Vascular Diseases | L94.0, L94.1, L94.3, M05.x, M06.x, M08.x, M12.0, M12.3, M30.x, M31.0-M31.3, M32.x-M35.x, M45.x, M46.1, M46.8, M46.9 |
| Coagulopathy | D65-D68.x, D69.1, D69.3-D69.6 |
| Obesity | E66.x |
| Weight Loss | E40.x-E46.x, R63.4, R64 |
| Fluid and Electrolyte Disorders | E22.2, E86.x, E87.x |
| Blood Loss Anemia | D50.0 |
| Deficiency Anemia | D50.8, D50.9, D51.x-D53.x |
| Alcohol Abuse | F10, E52, G62.1, I42.6, K29.2, K70.0, K70.3, K70.9, T51.x, Z50.2, Z71.4, Z72.1 |
| Drug Abuse | F11.x-F16.x, F18.x, F19.x, Z71.5, Z72.2 |
| Psychoses | F20.x, F22.x-F25.x, F28.x, F29.x, F30.2, F31.2, F31.5 |
| Depression | F20.4, F31.3-F31.5, F32.x, F33.x, F34.1, F41.2, F43 |

#### A.2. Distribution of Comorbidity Conditions by Race

Table S2 examines the demographic and clinical characteristics for COVID-19 patients treated at Michigan Medicine between March 10, 2020 - March 10, 2021, stratified by self-reported race. This was done to assess the patient mix by self-reported race group, to better understand how the distribution of comorbid conditions varies across these patient sub-populations.

Table S2: Demographic and clinical characteristics for the patient population, stratified by self-reported race

| **Characteristic** | **Black**  **N = 1,025^1^** | **Non-Black**  **N = 5,167^1^** | **Unknown**  **N = 539^1^** |
| --- | --- | --- | --- |
| Age (years) | 47 (31, 60) | 44 (24, 61) | 30 (19, 51) |
| Sex |  |  |  |
| Female | 609 (59%) | 2,876 (56%) | 221 (47%) |
| Male | 416(41%) | 2,290 (44%) | 246 (53%) |
| Body Mass Index (kg/m^2^) | 30 (29, 35) | 30(27, 30) | 30 (30, 30) |
| Congestive Heart Failure | 135 (13%) | 419 (8.1%) | 28 (5.2%) |
| Cardiac Arrhythmias | 373 (36%) | 1,373 (27%) | 51 (9.5%) |
| Valvular Disease | 63 (6.1%) | 324 (6.3%) | 4 (0.7%) |
| Pulmonary Circulation Disorders | 115 (11%) | 269 (5.2%) | 15 (2.8%) |
| Peripheral Vascular Disorders | 119 (12%) | 413 (8.0%) | 11 (2.0%) |
| Hypertension, Uncomplicated | 505 (49%) | 1,567 (30%) | 67 (12%) |
| Hypertension, Complicated | 192 (19%) | 418 (8.1%) | 17 (3.2%) |
| Paralysis | 30 (2.9%) | 105 (2.0%) | 3 (0.6%) |
| Other Neurological Disorders | 133 (13%) | 465 (9.0%) | 18 (3.3%) |
| Chronic Pulmonary Disease | 341 (33%) | 1,356 (26%) | 33 (6.1%) |
| Diabetes, Uncomplicated | 294 (29%) | 710 (14%) | 45 (8.3%) |
| Diabetes, Complicated | 226 (22%) | 482 (9.3%) | 35 (6.5%) |
| Hypothyroidism | 97 (9.5%) | 605 (12%) | 13 (2.4%) |
| Renal Failure | 235 (23%) | 481 (9.3%) | 28 (5.2%) |
| Liver Disease | 126 (12%) | 574 (11%) | 10 (1.9%) |
| Lymphoma | 22(2.1%) | 94 (1.8%) | 3 (0.6%) |
| Metastatic Cancer | 67 (6.5%) | 522 (10%) | 6 (1.1%) |
| Solid Tumor, Without Metastasis | 97 (9.5%) | 561 (11%) | 16 (3.0%) |
| Rheumatoid Arthritis/Collagen Vascular Diseases | 92 (9.0%) | 438 (8.5%) | 6 (1.1%) |
| Coagulopathy | 137 (13%) | 504 (9.8%) | 21 (3.9%) |
| Obesity | 447 (44%) | 1,396 (27%) | 37 (6.9%) |
| Weight Loss | 136 (13%) | 483 (9.3%) | 13 (2.4%) |
| Fluid and Electrolyte Disorders | 364 (36%) | 935 (18%) | 50 (9.3%) |
| Blood Loss Anemia | 84 (8.2%) | 187 (3.6%) | 5 (0.9%) |
| Deficiency Anemia | 180 (18%) | 470 (9.1%) | 10 (1.9%) |
| Alcohol Abuse | 48 (4.7%) | 229 (4.4%) | 8 (1.5%) |
| Drug Abuse | 78 (7.6%) | 253 (4.9%) | 5 (0.9%) |
| Psychoses | 51 (5.0%) | 145 (2.8%) | 5 (0.9%) |
| Depression | 318 (31%) | 1,489 (29%) | 28 (5.2%) |
| ^1^Median (IQR); n (%) |  |  |  |

### C. Fine-Gray Model for Competing Risks

Among hospitalized patients, we accounted for the competing relationship between two short-term outcomes of hospitalization, that is, discharge versus in-hospital death, using the Fine-Gray subdistribution regression model.^1^ By competing, we mean that in-hospital death and discharge censor each other. More specifically, let $\delta=1, 2$ index discharge and in-hospital death, respectively, and we observe either $\delta=1$ or $\delta=2$, but not both. Denote by $T$ the length of time from admission to discharge or death, whichever comes first. Then, for $j=1, 2$, define:

$F_{j}\left( t, \text{covariates} \right)=\Pr\left( T\leq t, \delta=j | \text{covariates} \right)$	and	$\lambda_{j}\left( t, \text{covariates} \right)=\frac{dF_{j}\left( t, \text{covariates} \right)/dt}{1 - F_{j}\left( t, \text{covariates} \right)}$

to be the covariate-dependent subdistribution function and subdistribution hazard function, respectively. The model in Fine and Gray (1999)^1^ stipulates that:

$$\lambda_{j}\left( t, \text{covariates} \right) = \lambda_{j0}\left( t \right)\exp\left( \beta_{j}\times\text{covariates} \right)$$

where $\lambda_{j0}\left( t \right)$ is unspecified for $j=1,2$. Therefore, the $\beta_{j}$ parameters have an interpretation of subdistribution hazard ratios for the outcomes of discharge $\left( j=1 \right)$ and in-hospital death $\left( j=2 \right)$, respectively. The subdistribution hazard ratio is an analog of hazard ratio, but in the context of competing risk models. The subdistribution hazard for a specific cause (say, discharge versus in-hospital death in our setting) is defined to be the probability of failure due to that cause at a moment in time, given that no failures of that cause have occurred thus far. The subdistribution hazard ratio is the ratio of subdistribution hazards corresponding to two conditions (e.g., having hypertension versus not).

### D. Additional Results

Additional model results are presented in this section. Namely, we fit two additional models to further study the outcomes of our study population: (1) hospitalization-free survival among all COVID-19 and (2) readmission post-index discharge among hospitalized patients.

#### C.1. Hospitalization-Free Survival

Table S3 presents the adjusted associations of patient demographic and clinical risk factors based on a Cox proportional hazards regression for hospitalization-free survival among all COVID-19 patients. Hospitalization-free survival was defined as the time from COVID-19 diagnosis to first admission date or death, whichever came first, subject to censoring by the end of this study. Older (per 10 years; HR: 1.23; 95% CI: 1.18-1.28), male (1.34; 1.16-1.54), and Black patients (1.89; 1.61-2.17) had higher hospitalization hazards. Additionally, ten comorbidity conditions were associated with higher hazards of hospitalization: cardiac arrhythmias (1.77; 1.51-2.07), pulmonary circulation disorders (1.27; 1.05-1.53), hypertension with (1.21; 1.01-1.45) and without complications (1.49; 1.22-1.83), other neurological disorders (1.35; 1.14-1.60), diabetes with complications (1.28; 1.02-1.60), coagulopathy (1.87; 1.60-2.20), weight loss (1.22; 1.03-1.44), fluid and electrolyte disorders (4.48; 3.78-5.30), and blood loss anemia (1.32; 1.06-1.64), while five were associated with lower hospitalizations hazards: valvular disease (0.71; 0.57-0.89), liver disease (0.79; 0.67-0.95), rheumatoid arthritis/collagen vascular diseases (0.80; 0.64-0.99), deficiency anemia (0.70; 0.58-0.84), and alcohol abuse (0.62; 0.46-0.83).

#### C.2. Readmissions

Studying the risk factors for readmission post-discharge, we found 256 that cardiac arrhythmias (Odds Ratio (OR): 1.85; 95% CI: 1.12-3.13), peripheral vascular disorders (1.66; 1.01-2.71), weight loss (1.69; 1.10-2.59), fluid and electrolyte disorders (1.84; 1.08, 3.22), blood loss anemia (1.81; 1.06-3.08), and depression (2.20; 1.42-3.42) were significantly associated with higher odds of readmission, while older age (per ten years) was associated with a 16% lower odds of readmission (0.84; 0.74-0.95) and chronic pulmonary disease had lower odds of readmission (0.63; 0.40-0.98; Table S4).

Table S3: Adjusted associations for hospitalization-free survival among all COVID-19 patients

| **Characteristic** | **Hospitalization-Free Survival**  **HR (95% CI)^1^** |
| --- | --- |
| Age (years) | **1.23 (1.18, 1.28)^2^** |
| Sex |  |
| Female | — |
| Male | **1.34 (1.16, 1.54)** |
| Race |  |
| Black | **—** |
| Non-Black | **0.53 (0.46, 0.62)** |
| Unknown | **0.73 (0.54, 1.00)** |
| Body Mass Index (kg/m^2^) | 1.01 (1.00, 1.02) |
| Congestive Heart Failure | 0.83 (0.69, 1.00) |
| Cardiac Arrhythmias | **1.77 (1.51, 2.07)** |
| Valvular Disease | **0.71 (0.57, 0.89)** |
| Pulmonary Circulation Disorders | **1.27 (1.05, 1.53** |
| Peripheral Vascular Disorders | **0.81 (0.67, 0.98)** |
| Hypertension, Uncomplicated | **1.21 (1.01, 1.45)** |
| Hypertension, Complicated | **1.49 (1.22, 1.83)** |
| Paralysis | 1.16 (0.88, 1.54) |
| Other Neurological Disorders | **1.35 (1.14, 1.60)** |
| Chronic Pulmonary Disease | 0.98 (0.84, 1.14) |
| Diabetes, Uncomplicated | 1.01 (0.82, 1.25) |
| Diabetes, Complicated | **1.28 (1.02, 1.60)** |
| Hypothyroidism | 0.93 (0.77, 1.12) |
| Renal Failure | 0.84 (0.70, 1.02) |
| Liver Disease | **0.79 (0.67, 0.95)** |
| Lymphoma | 0.81 (0.56, 1.17) |
| Metastatic Cancer | 0.82 (0.65, 1.04) |
| Solid Tumor, Without Metastasis | 0.92 (0.74, 1.15) |
| Rheumatoid Arthritis/Collagen Vascular Diseases | **0.80 (0.64, 0.99)** |
| Coagulopathy | **1.87 (1.60, 2.20)** |
| Obesity | 1.04 (0.89, 1.23) |
| Weight Loss | **1.22 (1.03, 1.44)** |
| Fluid and Electrolyte Disorders | **4.48 (3.78, 5.30)** |
| Blood Loss Anemia | **1.32 (1.06, 1.64)** |
| Deficiency Anemia | **0.70 (0.58, 0.84)** |
| Alcohol Abuse | **0.62 (0.46, 0.83)** |
| Drug Abuse | 1.12 (0.89, 1.42) |
| Psychoses | 0.92 (0.72, 1.18) |
| Depression | 0.88 (0.76, 1.02) |
| ^1^HR = Hazard Ratio; CI = Confidence Interval  ^2^Bold values indicate statistical significance |  |

Table S4: Adjusted associations for the odds of a readmission after index discharge among hospitalized patients

| **Characteristic** | **Readmission**  **OR (95% CI)^1^** |
| --- | --- |
| Age (years) | **0.84 (0.74, 0.95)** |
| Sex |  |
| Female | — |
| Male | 1.53 (0.99, 2.36) |
| Race |  |
| Black | — |
| Non-Black | 0.98 (0.63, 1.54) |
| Unknown | 0.22 (0.01, 1.12) |
| Body Mass Index (kg/m^2^) | 0.99 (0.96, 1.02) |
| Congestive Heart Failure | 0.87 (0.50, 1.49) |
| Cardiac Arrhythmias | **1.85 (1.12, 3.13)** |
| Valvular Disease | 1.22 (0.67, 2.19) |
| Pulmonary Circulation Disorders | 1.11 (0.67, 1.83) |
| Peripheral Vascular Disorders | **1.66 (1.01, 2.71)** |
| Hypertension, Uncomplicated | 1.31 (0.77, 2.26) |
| Hypertension, Complicated | 1.05 (0.56, 1.96) |
| Paralysis | 1.43 (0.73, 2.76) |
| Other Neurological Disorders | 1.31 (0.83, 2.07) |
| Chronic Pulmonary Disease | **0.63 (0.40, 0.98)** |
| Diabetes, Uncomplicated | 0.81 (0.42, 1.51) |
| Diabetes, Complicated | 1.03 (0.54, 1.98) |
| Hypothyroidism | 0.99 (0.57, 1.66) |
| Renal Failure | 0.92 (0.52, 1.63) |
| Liver Disease | 1.25 (0.78, 2.00) |
| Lymphoma | 0.49 (0.15, 1.39) |
| Metastatic Cancer | 1.65 (0.85, 3.18) |
| Solid Tumor, Without Metastasis | 1.46 (0.77, 2.72) |
| Rheumatoid Arthritis/Collagen Vascular Diseases | 1.42 (0.78, 2.53) |
| Coagulopathy | 1.41 (0.92, 2.17) |
| Obesity | 0.64 (0.38, 1.08) |
| Weight Loss | **1.69 (1.10, 2.59)** |
| Fluid and Electrolyte Disorders | **1.84 (1.08, 3.22)** |
| Blood Loss Anemia | **1.81 (1.06, 3.08)** |
| Deficiency Anemia | 1.25 (0.76, 2.05) |
| Alcohol Abuse | 1.01 (0.47, 2.08) |
| Drug Abuse | 0.99 (0.54, 1.79) |
| Psychoses | 1.04 (0.56, 1.89) |
| Depression | **2.20 (1.42, 3.42)** |
| ^1^OR = Odds Ratio; CI = Confidence Interval  ^2^Bold values indicate statistical significance |  |

### E. Additional Sensitivity Analysis Results

In a sensitivity analysis, we restricted hospitalizations to COVID-induced admissions, and only considered COVID-induced admission following discharge from a first COVID hospitalization. Table S5 reports the adjusted associations of the patient demographic and clinical risk factors with COVID-19 outcomes among patients hospitalized for COVID-19, based on a Fine-Gray subdistribution hazards regression for (a) discharge post-admission and (b) in-hospital death post-admission, and a logistic regression for (c) whether the patient was readmitted at any point post-index discharge.

#### D.1. Hospitalizations due to COVID-19

Among the COVID-induced hospitalizations, we modeled in-hospital death versus discharge using the Fine-Gray subdistribution hazards model. Consistent with the findings in the main text, older age (per 10 years) was associated with a 9% lower discharge rate (Subdistribution Hazard Ratio (SHR): 0.91; 95% CI: 0.86-0.96) and a 48% higher in-hospital mortality rate (SHR: 1.48; 95% CI: 1.25-1.75) post-diagnosis. Coagulopathy (Discharge: 0.71; 0.60-0.84; Mortality: 1.57; 1.04-2.37), fluid and electrolyte disorders (Discharge: 0.68; 0.57-0.80; Mortality: 2.54; 1.28-5.03), and blood loss anemia (Discharge: 0.67; 0.52-0.85; Mortality: 2.62; 1.62-4.23) were associated with lower discharge and higher inpatient mortality rates. Additionally, male sex (0.84; 0.72-0.98), diabetes with complications (0.71; 0.57-0.88), and other neurological disorders (0.72; 0.60-0.86) were associated with lower discharge rates, while chronic pulmonary disease (1.68; 1.12-2.52) was associated with higher inpatient mortality. In contrast, solid tumor cancers (1.31; 1.04-1.68), rheumatoid arthritis/collagen vascular diseases (1.37; 1.08-1.73), and drug abuse (1.34; 1.04-1.73) were associated with higher discharge rates, while weight loss (0.52; 0.31-0.89), deficiency anemia (0.46; 0.25-0.86) and depression (0.57; 0.34-0.98) were associated with lower inpatient mortality rates (Table S5).

#### D.2. Readmissions due to COVID-19

Studying the risk factors for COVID-induced readmission following the first COVID-induced hospitalization, we found that depression (2.61; 1.61-4.62) was significantly associated with higher odds of readmission, while chronic pulmonary disease (0.60; 0.36-0.98) patients had lower odds of readmission (Table S5). Owing to a smaller subset of patients and therefore less events, some previously significant associations could not be detected, namely cardiac arrhythmias, peripheral vascular disorders, weight loss, fluid and electrolyte disorders, and blood loss anemia, though the magnitudes of the point estimates remained consistent.

Table S5: Adjusted associations from sensitivity analysis among patients hospitalized for COVID-19

| **Characteristic** | **Discharge**  **SHR (95% CI)^1^** | **In-Hospital Death SHR (95% CI)^1^** | **Readmission**  **OR (95% CI)^1^** |
| --- | --- | --- | --- |
| Age (years) | **0.91 (0.86, 0.96)^2^** | **1.48 (1.25, 1.75)** | 0.94 (0.82, 1.07) |
| Sex |  |  |  |
| Female | — | — | — |
| Male | **0.84 (0.72, 0.98)** | 1.37 (0.85, 2.21) | 1.33 (0.83, 2.17) |
| Race |  |  |  |
| Black | — | — | — |
| Non-Black | 1.15 (0.99, 1.35) | 0.69 (0.43, 1.12) | 0.87 (0.54, 1.44) |
| Unknown | **0.48 (0.33, 0.72)** | **2.38 (1.21, 4.69)** | 0.25 (0.01, 1.25) |
| Body Mass Index (kg/m^2^) | 1.00 (0.99, 1.01) | 0.97 (0.94, 1.01) | 0.99 (0.96, 1.03) |
| Congestive Heart Failure | 0.87 (0.70, 1.08) | 1.32 (0.81, 2.17) | 0.96 (0.53, 1.71) |
| Cardiac Arrhythmias | 0.86 (0.73, 1.00) | 1.43 (0.83, 2.46) | 1.66 (0.95, 2.96) |
| Valvular Disease | 1.19 (0.92, 1.55) | 0.99 (0.55, 1.78) | 1.49 (0.79, 2.78) |
| Pulmonary Circulation Disorders | 0.86 (0.71, 1.04) | 1.01 (0.64, 1.61) | 1.30 (0.75, 2.22) |
| Peripheral Vascular Disorders | 1.19 (0.97, 1.47) | 0.75 (0.44, 1.26) | 1.03 (0.59, 1.77) |
| Hypertension, Uncomplicated | 1.02 (0.86, 1.21) | 0.72 (0.43, 1.20) | 1.38 (0.75, 2.57) |
| Hypertension, Complicated | 1.21 (0.98, 1.51) | 0.85 (0.48, 1.51) | 0.93 (0.47, 1.81) |
| Paralysis | 1.09 (0.83, 1.44) | 0.78 (0.30, 2.05) | 0.94 (0.43, 1.95) |
| Other Neurological Disorders | **0.72 (0.60, 0.86)** | 1.43 (0.94, 2.18) | 1.42 (0.86, 2.32) |
| Chronic Pulmonary Disease | **0.86 (0.73, 1.00)** | **1.68 (1.12, 2.52)** | **0.60 (0.36, 0.98)** |
| Diabetes, Uncomplicated | 1.10 (0.89, 1.35) | 0.91 (0.51, 1.61) | 0.75 (0.37, 1.49) |
| Diabetes, Complicated | **0.71 (0.57, 0.88)** | 1.79 (0.98, 3.26) | 0.98 (0.49, 2.00) |
| Hypothyroidism | 1.03 (0.84, 1.27) | 0.67 (0.33, 1.35) | 0.82 (0.45, 1.46) |
| Renal Failure | 0.87 (0.71, 1.07) | 1.13 (0.68, 1.87) | 1.12 (0.60, 2.09) |
| Liver Disease | 0.94 (0.77, 1.14) | 1.24 (0.77, 2.00) | 1.05 (0.62, 1.77) |
| Lymphoma | 0.68 (0.42, 1.09) | 1.66 (0.67, 4.11) | 0.78 (0.23, 2.26) |
| Metastatic Cancer | 1.16 (0.88, 1.52) | 0.98 (0.49, 1.96) | 1.55 (0.75, 3.16) |
| Solid Tumor, Without Metastasis | **1.32 (1.04, 1.68)** | 0.66 (0.34, 1.26) | 1.40 (0.71, 2.72) |
| Rheumatoid Arthritis/Collagen Vascular Diseases | **1.37 (1.08, 1.73)** | 0.77 (0.35, 1.72) | 1.02 (0.50, 1.97) |
| Coagulopathy | **0.71 (0.60, 0.84)** | **1.57 (1.04, 2.37)** | 1.27 (0.78, 2.05) |
| Obesity | 0.98 (0.81, 1.17) | 1.54 (0.94, 2.51) | 0.76 (0.43, 1.35) |
| Weight Loss | 0.91 (0.77, 1.07) | **0.52 (0.31, 0.89)** | 1.57 (0.98, 2.51) |
| Fluid and Electrolyte Disorders | **0.68 (0.57, 0.80)** | **2.54 (1.28, 5.03)** | 1.77 (0.97, 3.37) |
| Blood Loss Anemia | **0.67 (0.52, 0.85)** | **2.62 (1.62, 4.23)** | 1.72 (0.97, 3.03) |
| Deficiency Anemia | 1.04 (0.85, 1.27) | **0.46 (0.25, 0.86)** | 1.27 (0.73, 2.16) |
| Alcohol Abuse | 1.18 (0.85, 1.64) | 0.63 (0.28, 1.43) | 0.72 (0.29, 1.62) |
| Drug Abuse | **1.34 (1.04, 1.73)** | 0.85 (0.36, 2.04) | 1.30 (0.67, 2.47) |
| Psychoses | 1.03 (0.78, 1.37) | 1.12 (0.58, 2.16) | 0.90 (0.45, 1.73) |
| Depression | 1.07 (0.92, 1.24) | **0.57 (0.34, 0.98)** | **2.61 (1.61, 4.26)** |
| ^1^SHR = Subdistribution Hazard Ratio; CI = Confidence Interval; OR = Odds Ratio  ^2^Bold values indicate statistical significance | | | |

### F. Univariate Associations

Table S6: Univariate associations of clinical risk factors among all COVID-19 patients, adjusted for patient demographics

| **Characteristic** | **Number of Encounters**  **IRR (95% CI)^1,2^** | **Overall-Mortality**  **HR (95% CI)^1,2^** |
| --- | --- | --- |
| Body Mass Index (kg/m^2^) | **1.00 (1.00, 1.00)^3^** | 1.01 (0.98, 1.03) |
| Congestive Heart Failure | **2.26 (2.23, 2.29)** | **3.07 (2.20, 4.29)** |
| Cardiac Arrhythmias | **2.52 (2.50, 2.55)** | **4.51 (3.06, 6.65)** |
| Valvular Disease | **1.79 (1.76, 1.82)** | **1.77 (1.18, 2.66)** |
| Pulmonary Circulation Disorders | **2.61 (2.58, 2.64)** | **3.02 (2.11, 4.31)** |
| Peripheral Vascular Disorders | **2.30 (2.27, 2.32)** | **1.70 (1.18, 2.46)** |
| Hypertension, Uncomplicated | **2.08 (2.05, 2.10)** | **2.14 (1.40, 3.29)** |
| Hypertension, Complicated | **2.40 (2.37, 2.43)** | **4.53 (3.21, 6.41)** |
| Paralysis | **2.76 (2.71, 2.81)** | **2.26 (1.22, 4.20)** |
| Other Neurological Disorders | **2.03 (2.01, 2.06)** | **3.28 (2.35, 4.59)** |
| Chronic Pulmonary Disease | **1.69 (1.67, 1.70)** | **2.52 (1.82, 3.49)** |
| Diabetes, Uncomplicated | **1.77 (1.75, 1.79)** | **2.32 (1.67, 3.22)** |
| Diabetes, Complicated | **2.01 (1.99, 2.03)** | **3.62 (2.60, 5.03)** |
| Hypothyroidism | **1.66 (1.64, 1.68)** | 0.85 (0.53, 1.35) |
| Renal Failure | **2.15 (2.12, 2.17)** | **3.40 (2.40, 4.81)** |
| Liver Disease | **2.00 (1.97, 2.02)** | **1.85 (1.27, 2.70)** |
| Lymphoma | **1.92 (1.88, 1.97)** | 1.88 (0.96, 3.69) |
| Metastatic Cancer | **1.87 (1.84, 1.89)** | 1.24 (0.82, 1.86) |
| Solid Tumor without Metastasis | **1.62 (1.60, 1.64)** | 0.96 (0.64, 1.42) |
| Rheumatoid Arthritis/Collagen Vascular Diseases | **1.70 (1.67, 1.72)** | 1.04 (0.63, 1.74) |
| Coagulopathy | **2.54 (2.51, 2.56)** | **4.40 (3.18, 6.07)** |
| Obesity | **1.71 (1.70, 1.73)** | **1.83 (1.32, 2.54)** |
| Weight Loss | **2.40 (2.37, 2.42)** | **1.96 (1.36, 2.82)** |
| Fluid and Electrolyte Disorders | **2.87 (2.84, 2.89)** | **11.2 (7.09, 17.8)** |
| Blood Loss Anemia | **2.75 (2.71, 2.79)** | **5.09 (3.48, 7.45)** |
| Deficiency Anemia | **2.20 (2.18, 2.23)** | **1.57 (1.06, 2.33)** |
| Alcohol Abuse | **1.39 (1.37, 1.42)** | **1.83 (1.03,3.26)** |
| Drug Abuse | **2.14 (2.11, 2.17)** | 1.63 (0.90, 2.95) |
| Psychoses | **1.79 (1.76, 1.82)** | **1.88 (1.15, 3.07)** |
| Depression | **2.10 (2.08, 2.12)** | 1.28 (0.91, 1.81) |
| ^1^IRR = Incidence Rate Ratio, CI = Confidence Interval, HR = Hazard Ratio  ^2^Adjusted for age, sex, and race  ^3^Bold values indicate statistical significance | | |
